# Hepatitis B Virus (HBV) treatment eligibility in the UK: retrospective longitudinal cohort data to explore the impact of changes in clinical guidelines

**DOI:** 10.1101/2024.12.02.24318329

**Authors:** Cori Campbell, Tingyan Wang, Alexander J Stockdale, Stacy Todd, Jakub Jaworski, Ben Glampson, Dimitri Papadimitriou, Erik Mayer, Hizni Salih, Gail Roadknight, Stephanie Little, Theresa Noble, Kinga A Várnai, Cai Davis, Ashley I Heinson, Michael George, Florina Borca, Timothy Roberts, Baptiste B Ribeyre, Louise English, Leilei Zhu, NIHR HIC Viral Hepatitis and Liver Disease Consortium, Kerrie Woods, Jim Davies, Graham S Cooke, Eleni Nastouli, Salim I Khakoo, William Gelson, Ahmed Elsharkawy, Eleanor Barnes, Philippa C Matthews

## Abstract

**Objective:** Nucleos/tide analogue (NA) drugs are used for the long-term treatment of chronic hepatitis B virus (HBV) infection. In a landscape of changing clinical recommendations, we set out to quantify the prescription of NA drugs to date, and to determine the impact of relaxing treatment eligibility criteria in a unique large real-world dataset.

**Design:** We assimilated longitudinal data from adults with chronic HBV infection from six centres in England through the UK National Institute for Health Research (NIHR) Health Informatics Collaborative (HIC) viral hepatitis framework. We describe factors currently associated with receipt of NA treatment, and determine the proportion of the population who would become treatment eligible as thresholds change.

**Results:** We reviewed data for 7558 adults, with mean follow up of 4.0 years (SD 3.9 years). NA treatment was prescribed in 2014/7558 (26.6%), and in line with existing guidelines was associated with HBeAg positivity and ALT above the upper limit of normal (ULN). Treatment was significantly more likely in males, older adults, in Asian and Other ethnicities (as compared to White), and significantly less likely in socioeconomically deprived individuals. The proportion of individuals who were treatment eligible was 32.3% based on 2 records of ALT>ULN over 6-12 months; 41.7% based on ALT>ULN *and* VL > 2000 IU/ml; and 95.1% based on detectable VL *and* either ALT>ULN *or* age>30 years.

**Conclusion:** We quantify the proportion of the population living with HBV who may become treatment eligible as guidelines change, providing insights to support the implementation of clinical services.

**KEY MESSAGES (3-5 sentences required by *Gut*):** *• What is already known on this topic:* To date, only a minority of adults living with chronic hepatitis B (CHB) infection have been eligible for treatment with nucleos/tide analogue (NA) therapy. However, worldwide guidelines are changing, with recommendations for treatment of an increasing proportion of the population. There is a need for evidence to inform the design of services to meet the needs of people living with CHB as more of the population becomes treatment eligible.

*• What this study adds:* We have determined the proportion of the UK population of people living with HBV infection who are currently treated, and determined the increasing proportion who would become eligible as treatment criteria change, with this proportion reaching 95% based on the least stringent treatment thresholds.

*• How this study might affect research, practice or policy:* Our study provides crucial real-world evidence that can inform planning of service delivery and resource allocation for people living with HBV, in a landscape of changing clinical guidelines.

## INTRODUCTION

Chronic Hepatitis B (CHB) infection is a leading cause of morbidity from liver cancer and cirrhosis worldwide (1,2), ranking as the first and third leading cause of death from these conditions, respectively (3,4). The World Health Organization’s Global Health Sector Strategy on viral hepatitis identified key targets for the elimination of HBV as a public health issue by 2030, which include reducing incidence, morbidity and mortality, and expanding antiviral coverage to individuals eligible for treatment (5). The mainstay of antiviral treatment is administration of a nucleos/tide analogue (NA) agent, which supresses HBV replication through reversible inhibition of the viral reverse transcriptase enzyme (6). This reduces the long-term risks of cirrhosis and HCC. NA treatment typically comprises a once daily oral dose of tenofovir disoproxil fumarate (TDF) or entecavir (ETV). However, NA agents are not curative, and must be taken long term in most cases.

In the UK (and most European countries), CHB is typically managed in specialist secondary or tertiary care clinics (led by hepatology, gastroenterology, infectious diseases or sexual health services) (6,7). Laboratory investigations include HBV e-antigen (HBeAg) status, HBV DNA viral load (VL) quantification, co-infection status (HIV/HCV/HDV), hepatic transaminases (such as alanine transferase, ALT) (6). Liver fibrosis can be assessed by transient elastography, ultrasound, and/or by calculation of laboratory-based fibrosis scores. Therapy is currently reserved for those deemed at the highest risk of complications, based on algorithms that account for these variables, alongside age, sex, and family history, and potentially supported by liver biopsy if other results are equivocal or co-factors contributing to liver disease need to be assessed. On this basis, a minority of people living with HBV have met criteria for antiviral treatment to date (6,8,9), but for any individual, disease markers and treatment eligibility may change over time so regular surveillance is required.

In 2024, the WHO published a high-profile report highlighting that the global public health response for HBV is not on track (10), and in parallel released new global guidelines, relaxing HBV treatment criteria with the intent of simplifying treatment roll-out, reducing costs, and making treatment access more equitable, particularly for resource-limited settings (11). In parallel, some countries have updated their national recommendations, moving in the same direction towards wider treatment, exemplified by China (12) and Brazil (13). European and North American guideline committees (EASL and AASLD respectively) are currently rewriting clinical practice guidelines.

We set out to use electronic health record (EHR) data using the Health Informatics Collaborative (HIC) framework for viral hepatitis (14), which collects demographic and longitudinal clinical data held by secondary and tertiary care centres. Our objectives were to assess the proportion of who have already been prescribed NA agents, describe characteristics of this treated population and individual characteristics associated with antiviral treatment, and to determine the impact of potential changes to guidelines on the population who meet eligibility criteria for treatment.

## METHODS

### NIHR Health Informatics Collaborative (HIC)

The HIC collects individual-level anonymised longitudinal EHR data, as previously described (14,15). Data elements are defined according to the standardised NHS Data Dictionary (16). Parameters include demographics, laboratory tests, elastography scores, imaging, potential risk factors for liver disease and information regarding antiviral treatment. Socioeconomic data were available for a subset of five NHS Trusts; deprivation was ascertained according to Index of Multiple Deprivation (IMD) deciles, which were collapsed into quintiles for this analysis. Lengths of follow-up vary both by individual and by respective NHS site (due to variable clinical review and data collection processes). Ethnicity is defined according to census categories which are transferred into HIC coding.

Data storage and entry processes are unique to individual NHS sites, with different sites using a range of EHR interfaces (including Cerner, Millennium and Epic). The HIC Viral Hepatitis theme provides an informatics infrastructure via which data from all sites are harmonised and hosted on a central server, hosted by Oxford University Hospitals (OUH) NHS Foundation Trust. A comprehensive data governance framework has been developed to oversee data collection across the sites which use heterogenous EHR processes and environments. Approved researchers access data remotely via a secure Trusted Research Environment.

Viral Hepatitis HIC data are currently collected from ten participating NHS sites, which have geographical coverage across England. Data from six centres were included in analyses for this project, based on availability of antiviral HBV treatment data (Cambridge University Hospitals NHS Foundation Trust (CUH); Imperial College Healthcare NHS Trust (ICHT); Liverpool University Hospitals NHS Foundation Trust (LUH); OUH; University College London Hospitals NHS Foundation Trust (UCLH); University Hospital Southampton NHS Foundation Trust (UHS)).

### Ethics

The NIHR HIC Viral Hepatitis theme database was approved by South Central—Oxford C Research Ethics Committee (REF Number: 21/SC/0060). The requirement for written informed consent is waived because data have been anonymized before transmission to the central data repository, but individuals are able to opt out from having their data included via the National Data Opt-out (17). This specific project was approved by written submission to the NIHR HIC Viral Hepatitis steering group, with representation from all participating centres.

### Study design and follow-up

We retrospectively identified individuals with CHB in the NIHR HIC database, who were aged ≥18 years at time of earliest record of HBV diagnosis. Typically, CHB is defined as a persistence of HBsAg and/or HBV DNA for at least six months. However, as per terms agreed by the HIC viral hepatitis steering group, we used relaxed eligibility criteria, requiring only one positive HBsAg or DNA VL measurement, to avoid unnecessary exclusions. Data analysed here were collected between February 1997 and April 2023, but the period covered varies by site and by individual.

Treatment decisions are taken by clinicians in the centre leading care, and will typically be based on UK guidance (issued by NICE, The National Institute for Health and Care Excellence (6)) or European guidance (issued by EASL (9)), recognising that prescribing decisions are also influenced by individual clinical judgement and patient choice. We could not reliably explore time to treatment initiation as the time of first prescription is not robustly recorded, and a proportion of individuals are already receiving treatment at the time of entry to the cohort.

### Primary aims

Our primary outcome of interest was antiviral treatment status (defined as a binary parameter; 0 = no treatment record at any point during the period of observation; 1 = treatment recorded at any time point throughout the period of observation). We investigated how demographics and laboratory measurements were associated with antiviral treatment, and evaluated the impact of different treatment criteria on the proportion of the whole CHB population becoming eligible for NA therapy under new thresholds.

### Variable and outcome ascertainment

We considered different treatment scenarios, starting with the real-world observation of those already prescribed NA therapy, and calculating the proportion of individuals with a record of treatment to date, using this as a baseline estimate of eligibility. We next applied revised treatment criteria based on new and emerging clinical guidelines (Table 1) to determine how these would increase the proportion of those eligible for treatment. Categories are based on thresholds included in WHO guidelines and others (11–13,18), pending new guidelines from the major liver societies (AASLD, EASL, APASLD) and UK NICE recommendations. ALT above the upper limit of normal (ULN) was defined as ≥30 IU/ML in males and ≥19 IU/ML in females as per WHO guidelines (11). In each case we assume the originally treated population remains under treatment, while others become newly eligible and are therefore added to the baseline treated population.

**Table 1:**
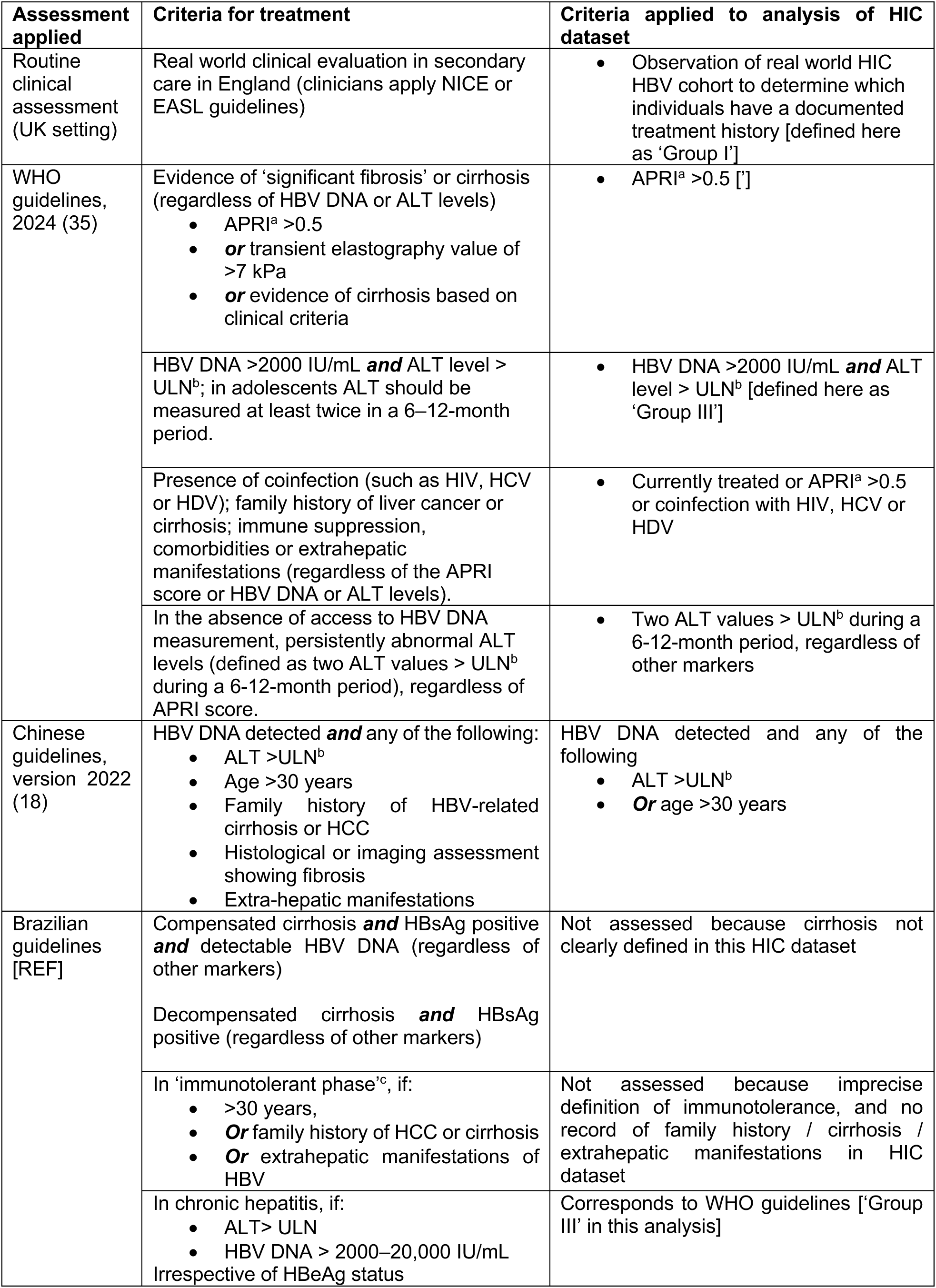
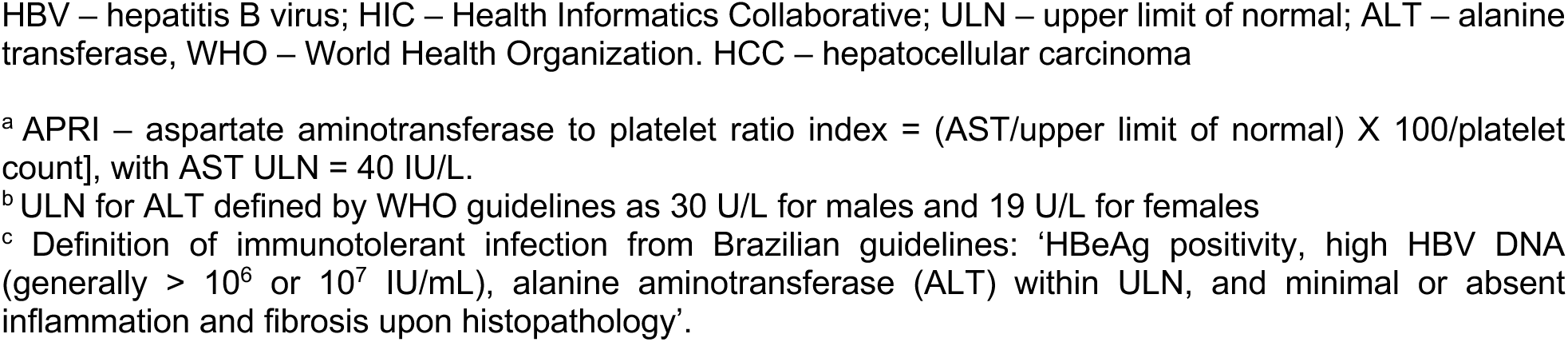
Thresholds applied to determine treatment eligibility in people living with chronic HBV infection, listed in the order of most to least stringent.

### Statistical analysis

We carried out analyses using R (version 4.1.0). Baseline characteristics were summarised for all CHB patients using descriptive statistics. Means and standard deviations (SDs) or medians and interquartile ranges (IQRs) were presented for continuous measures and were compared using *t* or Wilcoxon rank-sum tests, respectively. Patient counts and percentages were presented for continuous and binary variables; percentages were compared using chi-squared or Fisher’s exact tests.

Univariable and multivariable logistic regression models were used to investigate factors associated with antiviral treatment. Odds ratios (ORs) and 95% confidence intervals (95% CI) were reported for outputs. DNA VL and ALT were modelled as binary measures according to relevant thresholds (measurement > 2000 iu/mL and measurement > ULN, respectively).

### Data availability

The NIHR HIC Data Sharing Framework provides an overarching governance approach for data sharing between NHS organisations for research purposes. Research proposals are reviewed by the relevant NIHR HIC Scientific Steering Committees and in line with the NIHR HIC Data Sharing Framework. The governance terms of the HIC pertaining to clinical data for the viral hepatitis theme do not allow clinical data to be shared outside the Trusted Research Environment hosted by Oxford University Hospitals. More information about access to datasets within NIHR HIC data collaborations can be found on our website: https://hic.nihr.ac.uk/.

### Patient and public involvement statement

Our clinical teams work in close collaboration with patient representatives, who have been involved in planning prospective HIC studies.

## RESULTS

### Cohort characteristics at baseline

In total, 7558 adults living with HBV infection in England were eligible for inclusion in analysis (baseline characteristics in **Table 2**). Mean follow-up was 4.0 years (95% CI 3.9 to 4.1). Just under half the population were of White ethnicity (n = 3414, 45.2%), compared to 12.1% (n = 917) Asian, 17.8% (n = 1342) Black, 2.6% (n = 197) mixed, and 22.3% ‘other’ ethnicities (n = 1688). IMD was available for 6891 (91.2%) of our study population, among whom over half belonged to the two most deprived quintiles (most deprived IMD quintile (n = 1759, 25.5%) and second most deprived IMD quintile (1711, 24.8%)). A minority of individuals had documented coinfection with HIV (1.2%, n = 91), HCV (1.0%, n = 79) or HDV (1.0%, n = 75).

**Table 2:**
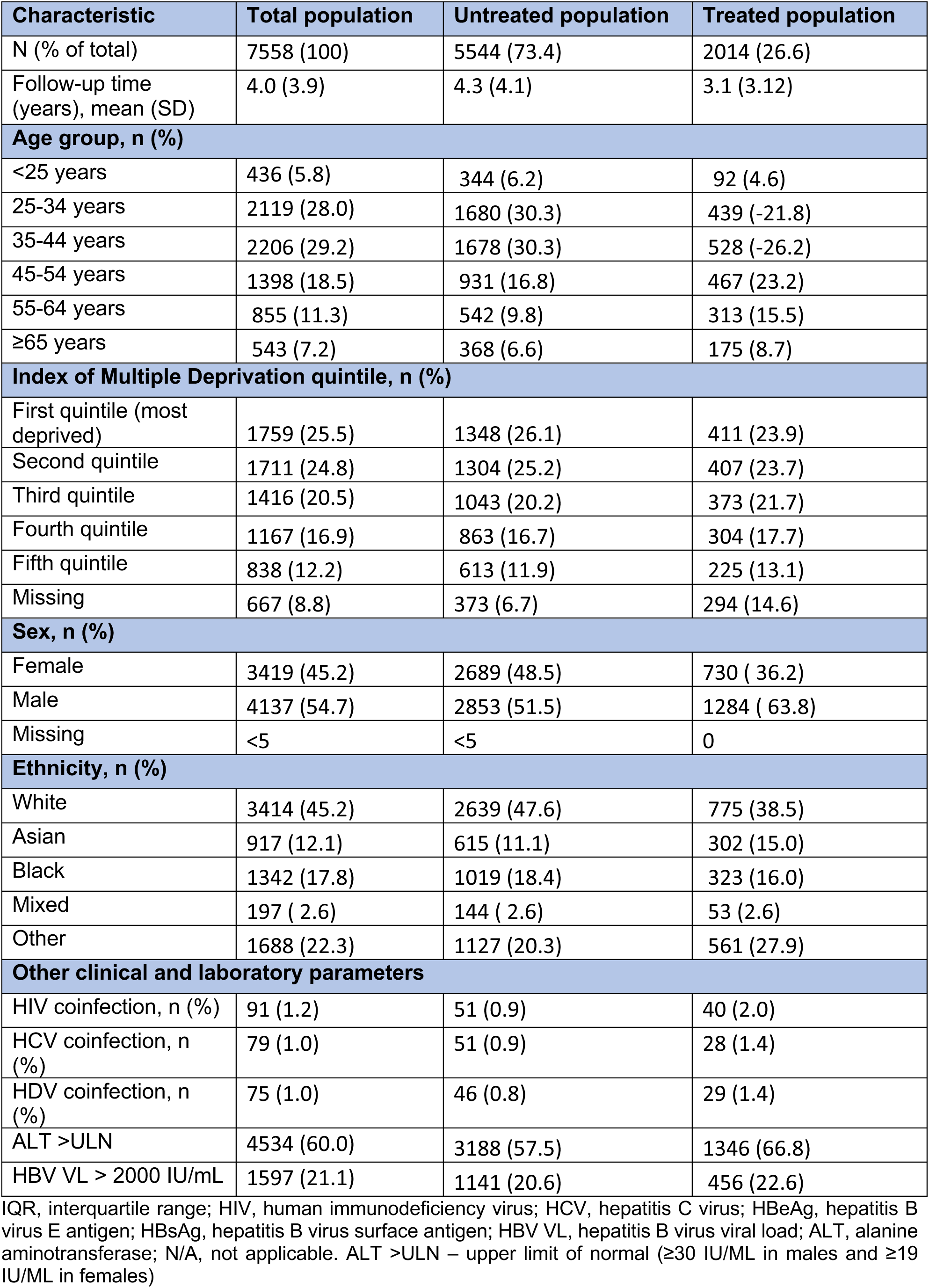
Baseline characteristics of individuals included in the NIHR HIC analysis investigating factors associated with antiviral treatment in chronic HBV infection. Percentages are expressed out of the column total, as stated in the first row.

| Characteristic | Total population | Untreated population | Treated population |
| --- | --- | --- | --- |
| N (% of total) | 7558 (100) | 5544 (73.4) | 2014 (26.6) |
| Follow-up time (years), mean (SD) | 4.0 (3.9) | 4.3 (4.1) | 3.1 (3.12) |
| **Age group, n (%)** |  |  |  |
| <25 years | 436 (5.8) | 344 (6.2) | 92 (4.6) |
| 25-34 years | 2119 (28.0) | 1680 (30.3) | 439 (-21.8) |
| 35-44 years | 2206 (29.2) | 1678 (30.3) | 528 (-26.2) |
| 45-54 years | 1398 (18.5) | 931 (16.8) | 467 (23.2) |
| 55-64 years | 855 (11.3) | 542 (9.8) | 313 (15.5) |
| ≥65 years | 543 (7.2) | 368 (6.6) | 175 (8.7) |
| **Index of Multiple Deprivation quintile, n (%)** |  |  |  |
| First quintile (most deprived) | 1759 (25.5) | 1348 (26.1) | 411 (23.9) |
| Second quintile | 1711 (24.8) | 1304 (25.2) | 407 (23.7) |
| Third quintile | 1416 (20.5) | 1043 (20.2) | 373 (21.7) |
| Fourth quintile | 1167 (16.9) | 863 (16.7) | 304 (17.7) |
| Fifth quintile | 838 (12.2) | 613 (11.9) | 225 (13.1) |
| Missing | 667 (8.8) | 373 (6.7) | 294 (14.6) |
| **Sex, n (%)** |  |  |  |
| Female | 3419 (45.2) | 2689 (48.5) | 730 (36.2) |
| Male | 4137 (54.7) | 2853 (51.5) | 1284 (63.8) |
| Missing | <5 | <5 | 0 |
| **Ethnicity, n (%)** |  |  |  |
| White | 3414 (45.2) | 2639 (47.6) | 775 (38.5) |
| Asian | 917 (12.1) | 615 (11.1) | 302 (15.0) |
| Black | 1342 (17.8) | 1019 (18.4) | 323 (16.0) |
| Mixed | 197 (2.6) | 144 (2.6) | 53 (2.6) |
| Other | 1688 (22.3) | 1127 (20.3) | 561 (27.9) |
| **Other clinical and laboratory parameters** |  |  |  |
| HIV coinfection, n (%) | 91 (1.2) | 51 (0.9) | 40 (2.0) |
| HCV coinfection, n (%) | 79 (1.0) | 51 (0.9) | 28 (1.4) |
| HDV coinfection, n (%) | 75 (1.0) | 46 (0.8) | 29 (1.4) |
| ALT >ULN | 4534 (60.0) | 3188 (57.5) | 1346 (66.8) |
| HBV VL > 2000 IU/mL | 1597 (21.1) | 1141 (20.6) | 456 (22.6) |

IQR, interquartile range; HIV, human immunodeficiency virus; HCV, hepatitis C virus; HBeAg, hepatitis B virus E antigen; HBsAg, hepatitis B virus surface antigen; HBV VL, hepatitis B virus viral load; ALT, alanine aminotransferase; N/A, not applicable. ALT >ULN – upper limit of normal (≥30 IU/ML in males and ≥19 IU/ML in females)

### Factors associated with receipt of antiviral treatment

A minority of the population had a record of antiviral treatment during the period of follow-up (n = 2014/7558, 26.6%; **Table 2)**. In the multivariable model, HBeAg positivity (OR 3.62, 95% CI 3.01 to 4.36) and ALT >ULN (OR 2.34, 95% CI 1.71 to 3.25) were associated with antiviral prescription (**Table 3**, **Figure 1)**, in line with clinical indications for starting treatment (6). Male sex (OR 1.67, 95% CI 1.44 to 1.96) and increasing age (ORs for ages 45-54 years, 55-64 years and ≥65 years age 1.77 (95% CI 1.43 to 2.20), 2.71 (95% CI 2.10 to 3.50) and 2.07 (95% CI 1.49 to 2.86), respectively) were also statistically associated with treatment (**Table 3**, **Figure 1)**. Individuals belonging to Asian and ‘Other’ ethnicities were more likely to be treated (ORs 1.76 (95% CI 1.40. to 2.20) and 1.71 (95% CI 1.42 to 2.04), respectively), compared to individuals of White ethnicity (**Table 3**, **Figure 1**). Belonging to the first IMD quintile (most deprived) was associated with a 24% reduction in the odds of antiviral treatment (OR 0.76, 95% CI 0.62 to 0.94) as compared to the third quintile.

**Figure 1.**
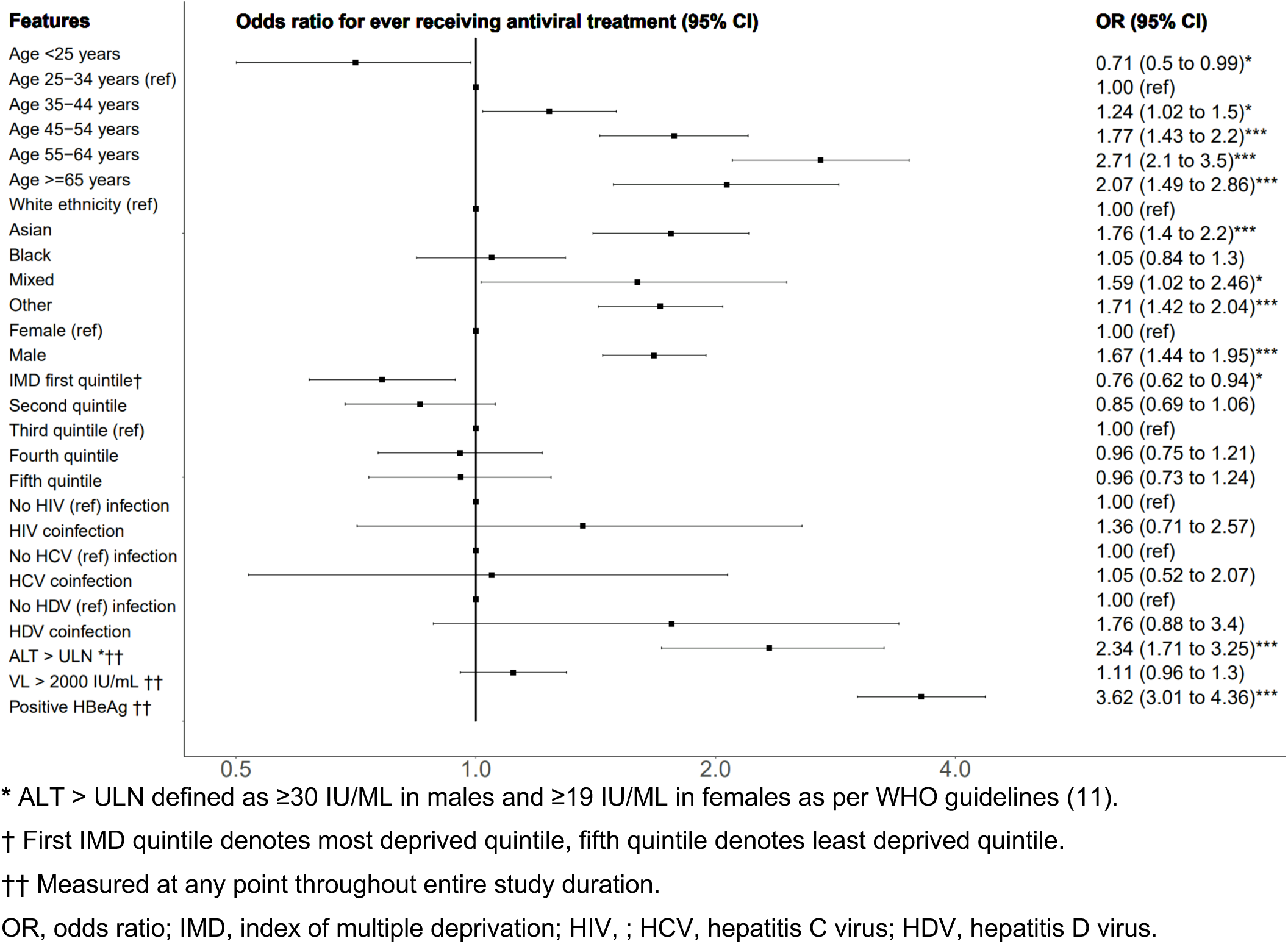
Forest plot showing factors associated with antiviral treatment in adults living with chronic Hepatitis B virus in the UK. Main multivariable logistic regression model from analysis undertaken in the NIHR Health Informatics Collaborative secondary care electronic health record database (n = 7558).

**Table 3.** Logistic regression models investigating factors which are associated with antiviral treatment in adults with CHB.

| Characteristic | Univariable OR (95% CI) | Multivariable OR (95% CI) |
| --- | --- | --- |
| **Age group** |  |  |
| <25 years | 1.02 (0.79 to 1.31) | 0.71 (0.50 to 0.99)* |
| 25-34 years | 1.00 (ref) | 1.00 (ref) |
| 35-44 years | 1.20 (1.04 to 1.39)* | 1.24 (1.02 to 1.50)* |
| 45-54 years | 1.92 (1.65 to 2.24)*** | 1.77 (1.43 to 2.20)*** |
| 55-64 years | 2.21 (1.86 to 2.63)*** | 2.71 (2.10 to 3.50)*** |
| ≥65 years | 1.82 (1.48 to 2.24)*** | 2.07 (1.49 to 2.86)*** |
| **Ethnicity** |  |  |
| White | 1.00 (ref) | 1.00 (ref) |
| Asian | 1.67 (1.42 to 1.96)*** | 1.76 (1.40 to 2.20)*** |
| Black | 1.08 (0.93 to 1.25) | 1.05 (0.84 to 1.30) |
| Mixed | 1.25 (0.9 to 1.72) | 1.59 (1.02 to 2.46)* |
| Other | 1.70 (1.49 to 1.93)*** | 1.71 (1.42 to 2.04)*** |
| **Sex** |  |  |
| Female | 1.00 (ref) | 1.00 (ref) |
| Male | 1.66 (1.49 to 1.84)*** | 1.67 (1.44 to 1.96)*** |
| **Index of Multiple Deprivation quintile** |  |  |
| First quintile (most deprived) | 0.85 (0.73 to 1.00) | 0.76 (0.62 to 0.94)* |
| Second quintile | 0.87 (0.74 to 1.03) | 0.85 (0.69 to 1.06) |
| Third quintile | 1.00 (ref) | 1.00 (ref) |
| Fourth quintile | 0.99 (0.83 to 1.17) | 0.96 (0.75 to 1.21) |
| Fifth quintile (least deprived) | 1.03 (0.85 to 1.24) | 0.96 (0.73 to 1.24) |
| **Coinfection status** |  |  |
| No HIV infection | 1.00 (ref) | 1.00 (ref) |
| HIV coinfection | 2.18 (1.43 to 3.31)*** | 1.36 (0.71 to 2.57) |
| No HCV infection | 1.00 (ref) | 1.00 (ref) |
| HCV coinfection | 1.52 (0.94 to 2.40) | 1.05 (0.52 to 2.07) |
| No HDV infection | 1.00 (ref) | 1.00 (ref) |
| HDV coinfection | 1.75 (1.08 to 2.77)* | 1.76 (0.88 to 3.40) |
| **Laboratory markers** |  |  |
| ALT > ULN<sup>†‡</sup> | 2.03 (1.65 to 2.52)*** | 2.34 (1.71 to 3.25)*** |
| HBV VL > 2000 IU/mL<sup>‡</sup> | 1.37 (1.22 to 1.53)*** | 1.11 (0.96 to 1.30) |
| HBeAg positive<sup>‡</sup> | 2.80 (2.43 to 3.22)*** | 3.62 (3.01 to 4.36)*** |

HIV, human immunodeficiency virus; HCV, hepatitis C virus; CT, computed tomography; ALT, alanine aminotransferase; HBV VL, hepatitis B virus viral load; HBeAg, hepatitis B virus E antigen.
\* $P < 0.05$
\*\* $P < 0.01$
\*\*\* $P < 0.001$
<sup>†</sup> ALT ULN (upper limit of normal) ≥30 IU/ML in males and ≥19 IU/ML in females
<sup>‡</sup> Mixed-effects logistic regression model, with unique individual identifier fitted as a random effect

### Influence of changing guidelines on proportion of population reaching treatment eligibility criteria

Having calculated the proportion of our clinical population with a record of antiviral treatment to date, we expanded this to reflect potential relaxation of treatment criteria (**Figure 2, Table S1**).

i. Observing the real-world cohort (in which therapy has been informed by UK or EASL guidelines (6,9) alongside patient and prescriber preference), **26.6%** (2014/7558) of individuals had a record of NA treatment.
ii. By adding an additional treatment criterion of two ALT measurements > ULN during any 6-12-month period, (regardless of other markers) an additional 726 individuals would have met treatment criteria during the period observed; thus overall **32.3%** (2740/7558) of the observed population would be offered treatment;
iii. Based on using an APRI score > 0.5 (suggesting liver fibrosis) as a treatment indicator in line with new WHO guidance (11), an additional 1123 individuals would have been considered treatment eligible, such that **41.5%** (3137/7558) of the observed population would be offered treatment. If accounting for the additional indication of HIV/HCV/HDV coinfection, an additional 111 patients become eligible, taking the total treated population to **43.0%** (3248/7558).
iv. Applying laboratory thresholds of HBV VL > 2000 IU/ml *and* ALT >ULN, in accordance with new WHO guidance (11) and with Brazilian national guidance (13), an additional 1138 individuals would meet criteria, such that **41.7%** (3152/7558) of the population would be offered treatment.
v. By applying updated Chinese treatment guidelines (18), which suggest offering treatment to all those with a detectable HBV VL with either ALT > ULN or age > 30 years, **95.1%** (n = 7187/7887) of individuals would potentially be treated;
vi. By definition, application of the most relaxed threshold would be to offer treatment to those with any marker of HBV infection (HBsAg and/or HBV DNA positivity), irrespective of quantification of other markers, making **100%** of the population eligible.

**Figure 2:**
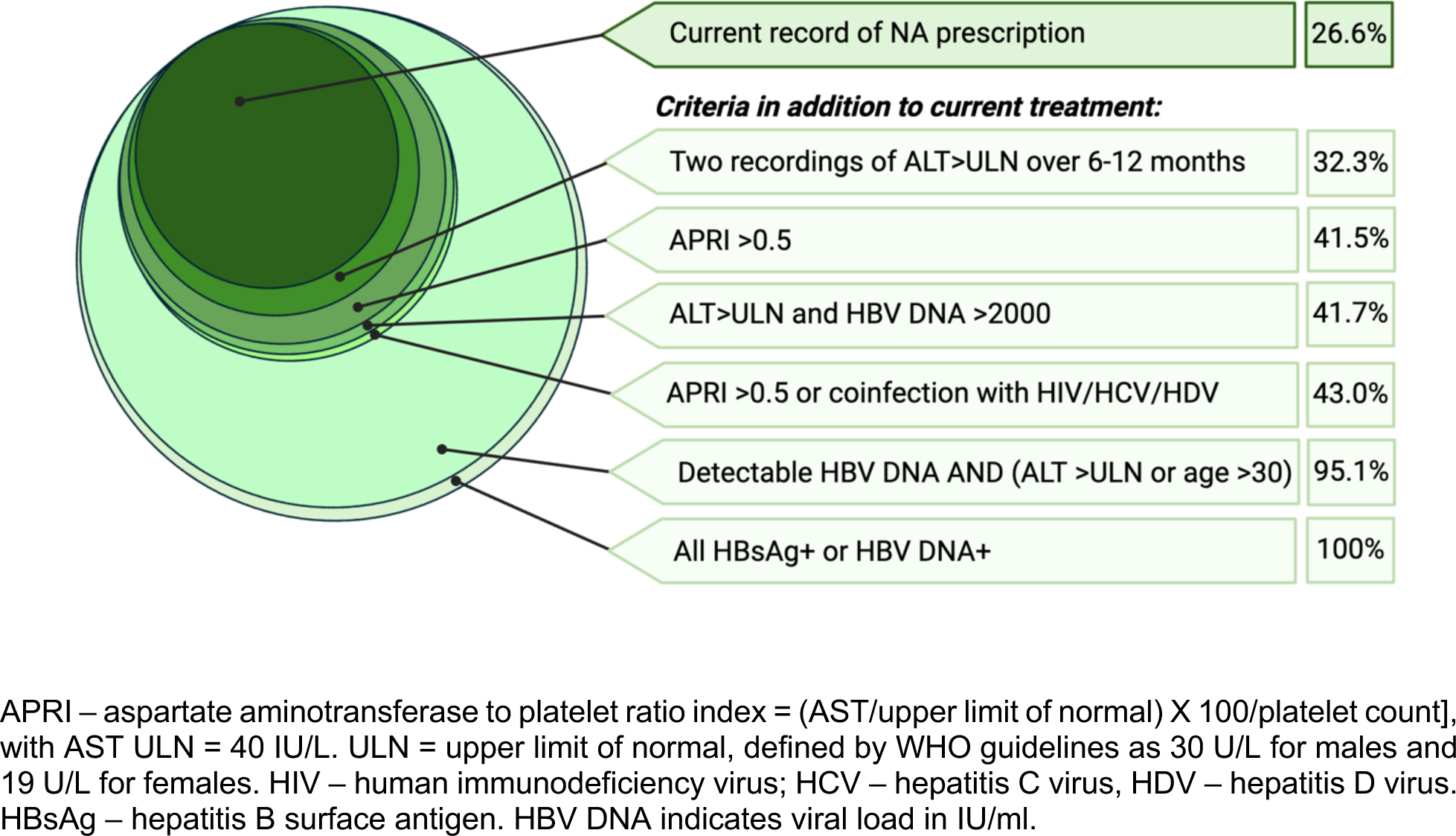
Visualisation of the impact of relaxing treatment criteria on the proportion of adults with CHB eligible for antiviral treatment with NA agents. We first calculated the proportion of the current population under treatment, and then considered six further scenarios, calculating the total currently on treatment plus those additionally meeting the relaxed treatment criteria in each of these. These data are also presented in tabulated form in Table S1.

In any of these scenarios, not all individuals offered antiviral therapy would wish to take it, and a small proportion may clear HBsAg (either in the context of acute or chronic infection), allowing therapy to be stopped. The estimates also do not account for individuals who might not meet laboratory criteria but would be offered treatment on other grounds, for example a biopsy result, clinical diagnosis of cirrhosis or family history of liver cancer, although these groups should be incorporated in those already on treatment at baseline in keeping with existing guidelines.

## DISCUSSION

### Antiviral treatment in CHB – current and future eligibility

In this analysis of adults with CHB accessing specialist secondary/tertiary care services across six centres in England, a minority (26.6%) had a record of NA treatment, reflecting recommendations in both UK/European (6,9) and global guidelines to date (8). This treated proportion is comparable to other cohorts (19–21). As treatment criteria are relaxed, an increasing proportion of the population will become treatment eligible, with our analysis suggesting that ∼30-50% will meet the relaxed laboratory thresholds that are most likely to be proposed.

Older guidelines, which inform prescribing practice to date, are based on large historical cohorts, often observing specific populations in which parameters at baseline were used to estimate the longer-term risk of liver-related events, (with a lack of high quality longitudinal off-treatment data). Previous recommendations have also been influenced by availability and affordability of drug therapy, potentially reflecting a need to ration resources in some settings. However, as safety, accessibility and affordability has improved (6), the WHO and other bodies are widening eligibility criteria to improve outcomes and reduce health inequities.

Expansion of treatment coverage would benefit individuals through reduction of risk of progression to end-stage disease, and brings public health benefits through reducing transmission and offering overall cost benefits (22,23). Thus, treatment expansion supports progression towards global 2030 elimination targets (5,24). However, the specific risks and benefits of treatment expansion vary between settings, and careful evaluation is required to assess optimum interventions in different populations.

### Factors associated with treatment eligibility

Although we found a relationship between treatment and HBeAg status and ALT >ULN, there was not a statistical relationship between HBV VL and receipt of treatment. This lack of association can be explained, as those who entered the cohort already on treatment will typically have suppressed VL throughout. We expected to find an association between HIV-status and receipt of treatment, since all those living with HIV should be treated, and regimens overlap with HBV therapy. However, the number in this group is small and we recognise some treatment data are missing for the HIV/HBV coinfection group in the HIC dataset, as a result of the shared care of HIV between clinical services; where antiviral prescriptions are issued through Sexual Health services, anonymisation of clinical records prevents complete linkage (accounting for an underestimate in the proportion of the HIV/HBV population prescribed treatment). NICE guidelines were last published in 2017, at which time treatment of people with HBV coinfected with either HCV and HDV was interferon-based; this landscape has dramatically changed with the advent of direct acting antivirals (DAA) for HCV and more recently the approval of bulevirtide for HDV. Prospective data collection will be needed to provide an updated view of treatment data for these populations, and as coinfection rates are highly variable between global settings, this epidemiology will have an influence on the proportion who are HBV-treatment eligible in different regions.

The association between ethnicity and antiviral treatment is likely to be influenced by unmeasured host and viral characteristics, including viral genotypes which influence clinical phenotype of infection, and cluster in different populations according to country of birth. The CHB population in the UK is disproportionately socioeconomically deprived (25), and it is likely that IMD quintile associates with diverse other factors that influence treatment access and eligibility. Further characterisation of the social and economic characteristics of the UK CHB population is warranted, with a need for special scrutiny of groups who are under-represented as a result of systematic barriers to care.

We recognise that in a minority of individuals there may be clinical rationale to stop therapy, particularly in the small minority of adults who clear HBsAg (26) and in the context of pregnancy in which prophylactic NA may be administered for a defined period peri-partum (27). This is not relevant to the overall research question addressed here, and for the purpose of this analysis we have assumed that the majority of adults who started antiviral treatment in the time period observed will have continued long-term, as recommended by NICE (6).

### Strengths and limitations

The HIC database is the largest secondary care data source containing longitudinal laboratory parameters for viral hepatitis in the UK and representing real-world clinical data collected from individuals who are accessing health services at multiple NHS Trusts across the country, with increasing potential for generalisation. However, the data presented here represent only six large, urban centres in England; trends may differ across settings and additional data will be required to determine the extent to which our observations apply in other regions. Furthermore, people living with HBV infection in the UK who are not attending routine secondary care services may have specific vulnerabilities (28) and potentially worse outcomes.

The HIC framework collects a subset of data, but does not systematically record other parameters that may influence treatment prescription such as comorbidity (e.g. metabolic liver disease, diabetes, immunosuppression), extrahepatic complications (e.g. glomerulonephritis, vasculitis), or family history. Likewise, as elastography scores have not been consistently recorded in different EPR systems, there is a high level of missingness (**Table S2**), and we were not able to investigate the association between elastography scores and treatment in this study.

Although we have the advantage of a diverse cohort representing a range of ethnicities (and therefore viral genotypes), the proportions of individuals who are treatment eligible will vary between settings, and further data are needed to represent other populations. Ethnicity is poorly captured in electronic health records, and country of birth may be a more relevant parameter to collect. The relationship between treatment eligibility and people reported as being in ‘other’ ethnic groups needs further scrutiny to understand the characteristics of individuals represented in this broad and non-specific category.

A future aspiration for the HIC is to focus more analysis on clinically important endpoints, focusing on cirrhosis and HCC, which will allow us to evaluate (i) these outcome markers as determinants of treatment eligibility (all patients in these groups should already be offered treatment) and (ii) how changes in prescribing practice influence the evolution of these outcomes. The updated dictionary for data collection now includes ICD-10 coding for endpoints, together with approaches for collecting data from free-text imaging reports using Natural Language Processing. At the time of this analysis, these data were not available for participating sites, although proof-of-principle has been established (29). Quantification of HBsAg is an additional variable that can potentially influence treatment eligibility, but has not been consistently measured in many centres to date.

### Landscape for treatment expansion

It is likely that national and international guidelines will follow the precedents set by the WHO, Brazil and China, based on consensus across expert bodies (30,31). The specific recommendations, thresholds and nuance of guidelines will undoubtedly remain – to some extent – setting dependent. While TDF and ETV are broadly efficacious, cheap, and well tolerated with minimal toxicity (6), prescription of these agents nevertheless requires access to diagnostics, laboratory testing, imaging, and a secure supply of affordable drugs. In regions where access to health infrastructure and laboratory stratification is challenging, the simplest assessments are required, including the option to offer treatment to any adult who tests HBsAg-positive. While HBV DNA assessment is widely available across the UK (and there are no additional costs to the patient to access this test), we have nevertheless applied WHO guidelines that account for situations in which this is not available in order to inform understanding of the potential impact of this simplified criterion. Expansion of treatment availability and eligibility may also require improved access to HBV DNA assays for the purpose of surveillance of resistance to antiviral agents (11,32).

Demands on health service infrastructure a key consideration to ensuring that treatment expansion can be implemented in the real world. In different settings, change in guidelines may accelerate decentralisation of HBV management to primary care, integration of HBV care with other services, and investment in triple elimination programmes that tackle HBV together with HIV and syphilis. Cost-effectiveness analysis of different models will be needed, supported by close partnerships between policymakers, clinical services and the pharmaceutical industry to support access to HBV therapy.

WHO guidelines now include treatment for adolescents (with those aged ≥12 years being treatment-eligible under the same criteria as adults). This group is outside the scope of this paper as HIC data only includes adults, but representation of younger people in research and clinical data will be crucial to understand implications for delivery of treatment.

### Patient representation

Patient voice and involvement of the community as stakeholders has increasingly been recognised as an important driver of guidance, recognising that individual preference is a fundamental component of therapeutic decision making (33,34), and with civil society organisations recognised as essential contributors to clinical guidelines. In many instances, those outside strict treatment criteria may express a wish to receive treatment, while in other cases those who do meet clinical thresholds for treatment may choose not to embark on life-long therapy.

### Conclusions

We have quantified treatment coverage across a large population of adults with CHB, demonstrating that a minority of individuals currently receive antiviral treatment but predicting the pattern of increasing eligibility in line with changing eligibility criteria. Such data are crucial to provide evidence to inform the resourcing, infrastructure and implementation of treatment programmes, and to tackle health inequities, thus supporting progress towards elimination targets.

## ACKNOWLEDGEMENTS

We would like to thank patients, clinicians, research nurses and research administrative staff at the contributing sites. The authors also extend their thanks to Deepak Suri and Stuart Flanagan from Department of Hepatology at UCLH who supported data collection. The authors thank Luca Mercuri from the Imperial BRC team, Karl McIntyre and Andrew Frankland from Liverpool Clinical Laboratories, as well as Luis Romão and David Ramlakhan, and Steve Harris from the UCLH BRC team, for their contributions on data collection.

## SUPPLEMENTARY MATERIAL

**Table S1.** Tabulation of how relaxing antiviral treatment initiation criteria would influence the number of individuals eligible for antiviral treatment. Seven different scenarios were considered (10).

| Scenario | Total, n (%) |
| --- | --- |
| (I) Currently receiving NA therapy, n (%) | 2014 (26.7) |
| (II) Scenario I AND/OR two ALT > ULN<sup>b</sup> during a 6-12-month period, regardless of other markers, n (%) | 2740 (32.3) |
| (III) Scenario I AND/OR APRI<sup>a</sup> >0.5 | 3137 (41.5) |
| (IV) Scenario I AND/OR ALT >ULN and VL > 2000 IU/ml, n (%) | 3152 (41.7) |
| (V) Scenario I AND/OR<br>• APRI > 0.5<br>OR<br>• Coinfected with HIV, HCV and/or HDV | 3248 (43.0) |
| (VI) Scenario I AND/OR Detectable VL AND:<br>• ALT > ULN<sup>b</sup><br>OR<br>• Age > 30 years | 7187 (95.1) |
| (VII) HBsAg and/or HBV DNA positivity (i.e. treatment of all individuals with markers of HBV infection) | 7558 (100) |

NA, nucleos(tide) analogue; ALT, alanine aminotransferase; VL, viral load; HBsAg, hepatitis B virus surface antigen, > ULN - Above the upper limit of normal (defined as ≥30 IU/ML in males and ≥19 IU/ML in females)
<sup>a</sup> APRI – aspartate aminotransferase to platelet ratio index = (AST/upper limit of normal) X 100/platelet count], with AST ULN = 40 IU/L.
<sup>b</sup> ULN for ALT defined by WHO guidelines as 30 U/L for males and 19 U/L for females.

**Table S2:** Types of hepatic imaging performed in individuals included in analysis of Health Informatics Collaborative data, investigating factors associated with antiviral treatment in chronic HBV infection. Data are presented from 3 NHS trusts contributing imaging data both overall (n = 2665) and stratified by those who had record of antiviral treatment (n = 547) and did not (2118) throughout follow-up. Percentages are expressed out of the column total.

| Characteristic | Total, n (%) | Untreated, n (%) | Treated, n (%) |
| --- | --- | --- | --- |
| N | 2665 | 2118 | 547 |
| Elastography, n (%) | 355 (13.3) | 336 (15.9) | 19 (3.5) |
| Liver stiffness score, mean (SD) | 6.23 (5.28) | 6.11 (5.21) | 8.33 (6.13) |
| MRI, n (%) | 65 (2.4) | 28 (1.3) | 37 (6.8) |
| CT, n (%) | 106 (4.0) | 61 (2.9) | 45 (8.2) |
| Ultrasound, n (%) | 1018 (38.2) | 632 (29.8) | 386 (70.6) |

SD, standard deviation; MRI, magnetic resonance imaging; CT, computed tomography.

## REFERENCES

1. World Health Organization. Hepatitis B Fact Sheet. [Internet]. 2022. Available from: https://www.who.int/en/news-room/fact-sheets/detail/hepatitis-b

2. Hepatitis B - FAQs, Statistics, Data, & Guidelines | CDC [Internet]. [cited 2023 Feb 16]. Available from: https://www.cdc.gov/hepatitis/hbv/index.htm

3. Fitzmaurice C, Akinyemiju T, Abera S, Ahmed M, Alam N, Alemayohu MA, et al. The burden of primary liver cancer and underlying etiologies from 1990 to 2015 at the global, regional, and national level results from the global burden of disease study 2015. JAMA Oncol [Internet]. 2017 Dec 1 [cited 2021 Apr 10];3(12):1683–91. Available from: /pmc/articles/PMC5824275/

4. Sheena BS, Hiebert L, Han H, Ippolito H, Abbasi-Kangevari M, Abbasi-Kangevari Z, et al. Global, regional, and national burden of hepatitis B, 1990–2019: a systematic analysis for the Global Burden of Disease Study 2019. Lancet Gastroenterol Hepatol [Internet]. 2022 Jun 1 [cited 2022 Aug 10];7(9):796–829. Available from: http://www.thelancet.com/article/S2468125322001248/fulltext

5. Global Health Sector Strategy on viral hepatitis 2016–2021. [Internet]. Geneva; 2016 [cited 2021 Jun 14]. Available from: https://apps.who.int/iris/bitstream/handle/10665/246177/WHO-HIV-2016.06-eng.pdf?sequence=1&isAllowed=y

6. National Institute for Health and Care Excellence. Hepatitis B (chronic): diagnosis and management (CG165). 2017;2016(June):1–36. Available from: https://www.nice.org.uk/guidance/cg165

7. Sarri G, Westby M, Bermingham S, Hill-Cawthorne G, Thomas H. Diagnosis and management of chronic hepatitis B in children, young people, and adults: summary of NICE guidance. BMJ [Internet]. 2013 [cited 2023 Apr 4];346:f3893. Available from: https://pubmed.ncbi.nlm.nih.gov/23804177/

8. Tan M, Bhadoria AS, Cui F, Tan A, Van Holten J, Easterbrook P, et al. Estimating the proportion of people with chronic hepatitis B virus infection eligible for hepatitis B antiviral treatment worldwide: a systematic review and meta-analysis. Lancet Gastroenterol Hepatol. 2021 Feb 1;6(2):106–19.

9. European Association for the Study of the Liver. EASL 2017 Clinical Practice Guidelines on the management of hepatitis B virus infection. J Hepatol. 2017;67:370–98.

10. World Health Organization. Global hepatitis report 2024: action for access in low- and middle-income countries [Internet]. 2024 [cited 2024 Jun 28]. Available from: https://www.who.int/publications/i/item/9789240091672

11. World Health Organization. Guidelines for the prevention, diagnosis, care and treatment for people with chronic hepatitis B infection. 2024;(March):1–272.

12. Guidelines for the prevention and treatment of chronic hepatitis B (version 2022). Zhonghua Gan Zang Bing Za Zhi [Internet]. 2022 Dec 20 [cited 2023 Sep 20];30(12):1309–31. Available from: https://pubmed.ncbi.nlm.nih.gov/36891718/

13. Ministério Da Saúde. HEPATITE B PROTOCOLO CLÍNICO E DIRETRIZES TERAPÊUTICAS DE E COINFECÇÕES. 2023 [cited 2024 Jun 28]; Available from: http://conitec.gov.br/

14. Wang T, Smith DA, Campbell C, Freeman O, Moysova Z, Noble T, et al. Cohort Profile: The National Institute for Health Research Health Informatics Collaborative: Hepatitis B Virus (NIHR HIC HBV) research dataset. Int J Epidemiol [Internet]. 2023 Feb 8 [cited 2023 May 7];52(1):e27–37. Available from: https://academic.oup.com/ije/article/52/1/e27/6609198

15. Smith DA, Wang T, Freeman O, Crichton C, Salih H, Matthews PC, et al. National Institute for Health Research Health Informatics Collaborative: development of a pipeline to collate electronic clinical data for viral hepatitis research. BMJ Health Care Inform [Internet]. 2020 Nov 1 [cited 2023 Jan 11];27(3):e100145. Available from: https://informatics.bmj.com/content/27/3/e100145

16. NHS Data Model and Dictionary [Internet]. [cited 2023 May 9]. Available from: https://www.datadictionary.nhs.uk/

17. National data opt-out - NHS Digital [Internet]. [cited 2023 May 9]. Available from: https://digital.nhs.uk/services/national-data-opt-out

18. You H, Wang F, Li T, Xu X, Sun Y, Nan Y, et al. Guidelines for the Prevention and Treatment of Chronic Hepatitis B (version 2022). http://www.xiahepublishing.com/ [Internet]. 2023 Nov 28 [cited 2024 Aug 26];11(6):1425–42. Available from: http://www.xiahepublishing.com/2310-8819/JCTH-2023-00320

19. Makuza JD, Jeong D, Wong S, Binka M, Adu PA, Velásquez García HA, et al. Association of hepatitis B virus treatment with all-cause and liver-related mortality among individuals with HBV and cirrhosis: a population-based cohort study. The Lancet Regional Health - Americas [Internet]. 2024 Aug 1 [cited 2024 Jun 30];36:100826. Available from: https://linkinghub.elsevier.com/retrieve/pii/S2667193X24001534

20. Davidov Y, Abu Baker F, Israel A, Ben Ari Z. Real-world hepatitis B antiviral treatment trends and adherence to practice guidelines: a large cohort study. Acta Clin Belg [Internet]. 2023 [cited 2024 Jun 30];78(4):291–7. Available from: https://pubmed.ncbi.nlm.nih.gov/36448668/

21. Ko KL, To WP, Mak LY, Seto WK, Ning Q, Fung J, et al. A large real-world cohort study examining the effects of long-term entecavir on hepatocellular carcinoma and HBsAg seroclearance. J Viral Hepat [Internet]. 2020 Apr 1 [cited 2024 Jun 30];27(4):397–406. Available from: https://onlinelibrary.wiley.com/doi/full/10.1111/jvh.13237

22. Chien RN;, Liaw YF, Bartosch B, Chien RN, Liaw YF. Current Trend in Antiviral Therapy for Chronic Hepatitis B. Viruses 2022, Vol 14, Page 434 [Internet]. 2022 Feb 21 [cited 2023 May 15];14(2):434. Available from: https://www.mdpi.com/1999-4915/14/2/434/htm

23. Zhang S, Wang C, Liu B, Lu Q Bin, Shang J, Zhou Y, et al. Cost-effectiveness of expanded antiviral treatment for chronic hepatitis B virus infection in China: an economic evaluation. Lancet Reg Health West Pac [Internet]. 2023 [cited 2023 May 15];0(0). Available from: http://www.thelancet.com/article/S2666606523000561/fulltext

24. Global health sector strategies on, respectively, HIV, viral hepatitis and sexually transmitted infections for the period 2022-2030.. Geneva; 2022.

25. Campbell C, Wang T, Gillespie I, Barnes E, Matthews PC. Characterisation of chronic hepatitis B virus infection in the UK and risk factors for hepatocellular carcinoma: a large electronic health record-based retrospective cohort study in the QResearch primary care database. medRxiv [Internet]. 2022 Sep 2 [cited 2022 Sep 7];2022.09.01.22279481. Available from: https://www.medrxiv.org/content/10.1101/2022.09.01.22279481v1

26. Downs LO, Smith DA, Lumley SF, Patel M, McNaughton AL, Mokaya J, et al. Electronic health informatics data to describe clearance dynamics of hepatitis B surface antigen (HBsAg) and e antigen (HBeAg) in chronic hepatitis B virus infection. mBio [Internet]. 2019 May 1 [cited 2024 Jun 30];10(3). Available from: https://journals.asm.org/doi/10.1128/mbio.00699-19

27. Matthews PC, Ocama P, Wang S, El-Sayed M, Turkova A, Ford D, et al. Enhancing interventions for prevention of mother-to-child-transmission of hepatitis B virus. JHEP Reports [Internet]. 2023 Aug 1 [cited 2024 Jun 30];5(8). Available from: /pmc/articles/PMC10405098/

28. Martyn E, Eisen S, Longley N, Harris P, Surey J, Norman J, et al. The forgotten people: Hepatitis B virus (HBV) infection as a priority for the inclusion health agenda. Elife. 2023 Feb 9;12.

29. Wang T, Glampson B, Mercuri L, Papadimitriou D, Jones CR, Smith DA, et al. Identifying Hepatocellular Carcinoma from imaging reports using natural language processing to facilitate data extraction from electronic patient records. medRxiv [Internet]. 2022 Aug 24 [cited 2022 Sep 23];2022.08.23.22279119. Available from: https://www.medrxiv.org/content/10.1101/2022.08.23.22279119v1

30. WHO announces the update of hepatitis B guidelines on testing and treatment [Internet]. [cited 2023 Sep 13]. Available from: https://www.who.int/news/item/29-04-2023-who-announces-the-update-of-hepatitis-b-guidelines-on-testing-and-treatment

31. Johannessen A. Is it time for a paradigm shift in treatment guidelines for chronic hepatitis B? Lancet Gastroenterol Hepatol [Internet]. 2023 Sep 1 [cited 2023 Sep 13];8(9):784. Available from: http://www.thelancet.com/article/S2468125323002364/fulltext

32. Goto A, Rodriguez-Esteban R, Scharf SH, Morris GM. Understanding the genetics of viral drug resistance by integrating clinical data and mining of the scientific literature. Scientific Reports 2022 12:1 [Internet]. 2022 Aug 25 [cited 2024 Dec 1];12(1):1–11. Available from: https://www.nature.com/articles/s41598-022-17746-3

33. Matthews PC, Jack K, Wang S, Abbott J, Bryce K, Cheng B, et al. A call for advocacy and patient voice to eliminate hepatitis B virus infection. Lancet Gastroenterol Hepatol [Internet]. 2022 Apr 1 [cited 2024 Jun 30];7(4):282–5. Available from: https://pubmed.ncbi.nlm.nih.gov/35278394/

34. Freeland C, Adjei C, Wallace J, Wang S, Hicks J, Adda D, et al. Survey of lived experiences and challenges in hepatitis B management and treatment. BMC Public Health [Internet]. 2024 Dec 1 [cited 2024 Jun 30];24(1):1–9. Available from: https://bmcpublichealth.biomedcentral.com/articles/10.1186/s12889-024-18425-w

35. World Health Organization. Guidelines for the prevention, diagnosis, care and treatment for people with chronic hepatitis B infection. 2024;

